# Trends in STI testing, diagnoses, and use of online chlamydia self-sampling services among young people during the first year of the COVID-19 pandemic in England

**DOI:** 10.1101/2023.03.22.23287571

**Authors:** Tamilore Sonubi, Dahir Sheik-Mohamud, Natasha Ratna, James Bell, Alireza Talebi, Catherine H Mercer, Katy Sinka, Stephanie J Migchelsen, Kate Folkard, Hamish Mohammed

**Affiliations:** Blood Safety, Hepatitis, Sexually Transmitted Infections (STI) and HIV Division – UK Health Security Agency, London, UK; Institute for Global Health, University College London, London, UK; The National Institute for Health Research Health Protection Research Unit in Blood Borne and Sexually Transmitted Infections at University College London in partnership with UK Health Security Agency, London, UK

**Keywords:** Young people, COVID-19 pandemic, STIs, Chlamydia, Gonorrhoea, Syphilis, Socioeconomic deprivation, online STI testing, COVID-19 impact, STI service provision

## Abstract

**Purpose:** Measures to control COVID-19 reduced face-to-face appointments and walk-ins at sexual health services (SHSs). Remote access to SHSs through online self-sampling for STIs was increased. This analysis assesses how these changes affected service use and STI testing among young people in England.

**Methods:** Data on all chlamydia, gonorrhoea and syphilis tests from 2019-2020 amongst English-resident 15-24 year olds (hereafter referred to as ‘young people’) were obtained from national STI surveillance datasets. We calculated proportional differences in tests and diagnoses for each STI, by demographic characteristics including age and socioeconomic deprivation, between 2019 and 2020. Among those tested for chlamydia, we used binary logistic regression to determine crude and adjusted odds ratios (OR) between demographic characteristics and being tested for chlamydia by an online service.

**Results:** Compared to 2019, there were declines in testing (30% for chlamydia, 26% for gonorrhoea, 36% for syphilis) and diagnoses (31% for chlamydia, 25% for gonorrhoea and 23% for syphilis) among young people in 2020. These reductions were greater amongst 15-19 year-olds (*vs*. 20-24 year-olds). Among young people tested for chlamydia, those living in the least deprived areas were more likely to be tested using an online self-sampling kit compared to those living in the most deprived areas (males; OR=1.24[1.22-1.26], females; OR=1.28[1.27-1.30]).

**Conclusion:** The first year of the COVID-19 pandemic in England saw declines in STI testing and diagnoses in young people and disparities in the use of online chlamydia self-sampling which risk widening existing health inequalities.

**Implications and contributions:** There was a decrease in STI testing of young people during the first year of the COVID-19 pandemic in England with larger reductions among teenagers. There was an increase in use of online STI self-sampling services but with inequalities in provision which risk widening existing inequalities in sexual health.

## Background

Sexually transmitted infections (STIs) present a public health challenge in England. The burden of STIs is greatest amongst young people aged 16-24 years and they are most likely to access sexual health services (SHSs) ^[2-6]^. Between 2010-12, they had the highest prevalence of chlamydia (3.1% of women and 2.3% of men in this age group). The prevalence of gonorrhoea was highest amongst people 20-24 years of age^[7]^.

The majority of chlamydia and gonorrhoea infections are asymptomatic, particularly amongst women, and may result in poor sexual and reproductive health if left untreated^[8]^; this includes a higher risk of pelvic inflammatory disease (PID), and ectopic pregnancy, which are preventable with early diagnosis and treatment^[9, 10]^. Consequently, opportunistic chlamydia screening has been offered to sexually active 15-24 year olds in England since 2003 through the National Chlamydia Screening Programme (NCSP).

There have been increases in attendances at SHSs between 2016-2020^[3]^. However, SHS delivery was greatly disrupted in March 2020 after the introduction of public health measures to reduce COVID-19 transmission such as national lockdowns, the requirement to stay at home, and social distancing. Moreover, the introduction of legislation restricting social interaction may have resulted in some people with STI-related symptoms avoiding attending SHSs due to fear of being judged for breaking these rules, creating a false sense of reduced demand on STI testing services^[11]^.

However, to ensure continued provision of STI testing, SHSs across England were rapidly reconfigured to provide more remote care via online consultations^[3]^.

To understand how STI testing, diagnoses and service use among 15-24 year olds in England changed during the first year of the COVID-19 pandemic, we compared the demographic characteristics of young people tested for STIs in 2019 and 2020 then, among all young people tested for chlamydia, determined the correlates of being tested via an online service instead of a face to face appointment.

## Methods

### Data description

We conducted a retrospective cohort study of 15-24 year olds (hereafter referred to as ‘young people’) residing in England who received an STI test or diagnosis utilising data from the GUMCAD STI Surveillance System and the CTAD Chlamydia Surveillance System. GUMCAD is a pseudonymised and depersonalised dataset of all attendances at SHSs in England and was used to obtain data on chlamydia, gonorrhoea and syphilis tests and diagnoses in young people attending this setting; syphilis diagnoses included primary, secondary and early latent stages^[12, 13]^. CTAD is a pseudonymised and depersonalised dataset of all publicly provided chlamydia tests and diagnoses, including those made through the NCSP, and was the source of data for chlamydia tests and diagnoses from community-based settings (those offering non-specialist STI-related care such as general practices and pharmacies)^[14, 15]^. All tests and diagnoses are coded by healthcare practitioners in keeping with surveillance reporting specifications. To avoid double counting of tests or diagnoses in each surveillance system, only one test or diagnosis of each STI for each person with a unique person identifier was counted within a 6-week episode^[15]^. Neither GUMCAD nor CTAD include personal identifiers so individuals cannot be matched between datasets; individuals are identified using a clinic-specific patient identification code in GUMCAD^[16]^ and a unique patient identifier number in CTAD^[14]^.

The study period was from 1^st^ January 2019 to 31^st^ December 2020 (inclusive), where data from 2019 relates to the pre-COVID period and 2020 relates to the first year of the COVID-19 pandemic. To be considered in the analysis data were restricted to people aged 15-24 years, at the time of the test or diagnosis, residing in England. Residential location was defined using the lower super output area (LSOA), small geographical areas for the reporting of small area statistics with an average size of 1,620 residents^[17]^. To obtain a measure of area-level socioeconomic deprivation, LSOAs were used to match to quintiles of the 2019 Index of Multiple Deprivation (IMD) dataset^[18]^. Additionally, the LSOAs were matched to the 2011 census area classification to categorise young people as living either in an urban or rural setting ^[19-21]^. Ethnicity was categorised using the national Census classification, as follows: Asian (including Bangladeshi, Chinese, Indian, Pakistani and any other Asian background), Black African, Black Caribbean, Other Black ethnicity, Mixed ethnicity (including White and Black Caribbean, White and Black African, White and Asian and any other Mixed or Multiple ethnic background), Other, and White^[22]^.

### Statistical analysis

We determined the proportional change in the characteristics of young people tested for STIs between 2019 and 2020. Demographic and clinical characteristics included age group (15-19 or 20-24 years), area of residence (rural or urban), residential area-level deprivation, (as defined by IMD quintile, where quintile 1 is the most deprived and quintile 5 is the least deprived), ethnicity, gender and public health region of residence (categorised as: East Midlands, East of England, London, North-East, North-West, South-East, South-West, West Midlands and Yorkshire and Humber) these characteristics were compared for all three STIs as they can be assessed in both CTAD and GUMCAD surveillance systems. Sexual orientation (including heterosexual males, men who have sex with men [MSM], heterosexual females and women who have sex with women [WSW]), and HIV status (categorised as; HIV diagnosed, HIV undiagnosed or unknown) were compared for gonorrhoea and syphilis as they are only collected in the GUMCAD surveillance system. Testing services (categorised as physical or online services) were compared for chlamydia only, as the CTAD surveillance system comprehensively captures all chlamydia testing from all publicly-commissioned testing services. The Pearson’s chi-square test was used to compare these characteristics across both years.

Subsequently, to assess any inequalities in the access to online self-sampling services for chlamydia testing (hereafter: ‘online chlamydia testing’), we restricted the sample to young people tested for chlamydia then used binary logistic regression to determine the crude and adjusted associations with being tested via an online service (yes vs. no); all models were stratified by gender. Bivariate models were created to determine the unadjusted odds ratios (ORs) for being tested online and residential area-level deprivation, as defined by IMD quintile (the primary independent variable), and each potential confounder (year of test, age group, area of residence and region of residence). All associations with a p-value less than 0.05 were considered to be statistically significant and all variables that had significant crude associations were included in the multivariable model. Adjusted odds ratios (aORs) were calculated using hierarchical modelling and covariates were added using a forward building approach. Firstly, Model 1 was constructed with year of test included a priori due to the scale up of online service provision during 2020^[3]^. The remaining covariates were added sequentially as follows: Model 2 was based on Model 1 with age group included as a confounder. Model 3 was based on Model 2 with the addition of area of residence. Lastly, Model 4 comprised Model 3 with the inclusion of region of residence. Ethnicity was excluded from the regression analysis due to a high degree of item non-response: 29% of young people tested for chlamydia were reported with an unspecified ethnic group in CTAD. All data analyses were performed using version Stata v15 (College Station, TX, USA)^[23]^.

## Results

### Trends in STI tests and diagnoses

There were 26-36% decreases in tests and diagnoses for chlamydia, gonorrhoea and syphilis among young people between 2019 and 2020 (Tables 1-3). However, there were greater proportional decreases among 15-19 compared to 20-24 year olds. By ethnicity, testing and diagnoses of all 3 STIs decreased for all ethnic groups with larger proportional declines among young people of Asian and Black non-African/non-Caribbean ethnicities. The number of chlamydia tests fell across all the different types of services offering testing (47%; 1,041,553 to 554,299), with the exception of online services where there was a 33% increase in testing between 2019 (271,684 tests) and 2020 (361,622 tests). Comparisons by sexual orientation could only be done for gonorrhoea and syphilis and, in both cases, testing and diagnoses fell to the largest extent (33-46%) among heterosexual men.

**Table 1.** Number of gonorrhoea test and diagnoses among 15–24 year olds residing in England, by demographic characteristics: 2019 to 2020.

|  | Tests |  |  | Diagnoses |  |  |
| --- | --- | --- | --- | --- | --- | --- |
|  | 2019 | 2020 | Percentage difference | 2019 | 2020 | Percentage difference |
| **Age** |  |  |  |  |  |  |
| 15 to 19 | 238,554 | 157,303 | -34% | 8,278 | 5,815 | -30% |
| 20 to 24 | 559,757 | 433,457 | -23% | 17,397 | 13,447 | -23% |
| **Ethnicity** |  |  |  |  |  |  |
| Asian | 31,993 | 20,516 | -36% | 985 | 641 | -35% |
| Black African | 35,797 | 26,257 | -27% | 1,396 | 1,259 | -10% |
| Black Caribbean | 29,370 | 21,071 | -28% | 1,892 | 1,345 | -29% |
| Other Black ethnicity | 9,130 | 5,755 | -37% | 513 | 353 | -31% |
| Mixed ethnicity | 47,461 | 35,606 | -25% | 2,175 | 1,729 | -21% |
| White | 554,275 | 382,189 | -31% | 16,393 | 11,327 | -31% |
| Other | 9,572 | 6,292 | -34% | 409 | 303 | -26% |
| Unknown ethnicity | 80,713 | 93,074 | 15% | 1,912 | 2,305 | 21% |
| **Sexual orientation** |  |  |  |  |  |  |
| Heterosexual Males | 196,172 | 131,192 | -33% | 6,557 | 4,554 | -31% |
| MSM | 47,550 | 39,356 | -17% | 6,525 | 4,782 | -27% |
| Heterosexual Females | 465,906 | 363,502 | -22% | 11,037 | 8,419 | -24% |
| WSW | 4,561 | 5,244 | 15% | 61 | 78 | 28% |
| Unknown sexual orientation | 84,122 | 51,466 | -39% | 1,495 | 1,429 | -4% |
| **Area of residence** |  |  |  |  |  |  |
| Rural | 79,697 | 60,021 | -25% | 1,590 | 1,100 | -31% |
| Urban | 704,883 | 518,388 | -26% | 23,643 | 17,773 | -25% |
| Unknown area of residence | 13,731 | 12,351 | -10% | 442 | 389 | -12% |
| **Residential area-level deprivation (Deprivation Quintile)\*** |  |  |  |  |  |  |
| 1 (most deprived) | 179,282 | 130,529 | -27% | 7,756 | 5,944 | -23% |
| 2 | 193,177 | 144,229 | -25% | 7,027 | 5,463 | -22% |
| 3 | 157,740 | 118,779 | -25% | 4,651 | 3,437 | -26% |
| 4 | 134,641 | 98,672 | -27% | 3,296 | 2,323 | -30% |
| 5 (least deprived) | 119,740 | 86,200 | -28% | 2,503 | 1,706 | -32% |
| Unknown deprivation quintile | 13,731 | 12,351 | -10% | 442 | 389 | -12% |
| **Region of residence** |  |  |  |  |  |  |
| East Midlands | 57,710 | 43,033 | -25% | 2,102 | 1,570 | -25% |
| East of England | 74,590 | 58,536 | -22% | 1,851 | 1,535 | -17% |
| London | 204,435 | 151,651 | -26% | 8,554 | 6,676 | -22% |
| North–East | 30,391 | 19,930 | -34% | 965 | 713 | -26% |
| North–West | 88,276 | 54,658 | -38% | 3,094 | 1,915 | -38% |
| South–East | 122,652 | 86,955 | -29% | 2,509 | 1,620 | -35% |
| South–West | 69,963 | 56,043 | -20% | 1,314 | 862 | -34% |
| West Midlands | 80,152 | 59,789 | -25% | 2,788 | 2,341 | -16% |
| Yorkshire and Humber | 59,577 | 48,957 | -18% | 2,144 | 1,676 | -22% |
| Unknown region of residence | 10,565 | 11,208 | 6% | 354 | 354 | 0% |
| **HIV status** |  |  |  |  |  |  |
| HIV diagnosed | 1,574 | 969 | -38% | 316 | 198 | -37% |
| HIV negative or unknown | 796,737 | 589,791 | -26% | 25,359 | 19,064 | -25% |
| **Total** | **798,311** | **590,760** | **-26%** | **25,675** | **19,262** | **-25%** |

\*Deprivation quintile is an area-level measure of deprivation and socioeconomic status based on the 2019 Index of Multiple Deprivation (IMD) quintiles

**Table 2.** Number of syphilis test and diagnoses among 15–24 year olds residing in England, by demographic characteristics: 2019 to 2020.

|  | Tests |  |  | Diagnoses |  |  |
| --- | --- | --- | --- | --- | --- | --- |
|  | 2019 | 2020 | Percentage difference | 2019 | 2020 | Percentage difference |
| **Age** |  |  |  |  |  |  |
| 15 to 19 | 136,686 | 78,620 | -42% | 223 | 139 | -38% |
| 20 to 24 | 382,761 | 255,340 | -33% | 948 | 766 | -19% |
| **Ethnicity** |  |  |  |  |  |  |
| Asian | 24,157 | 14,354 | -41% | 61 | 40 | -34% |
| Black African | 25,799 | 17,443 | -32% | 19 | 24 | 26% |
| Black Caribbean | 19,560 | 12,608 | -36% | 29 | 36 | 24% |
| Other Black ethnicity | 6,165 | 3,547 | -42% | 17 | 9 | -47% |
| Mixed ethnicity | 31,476 | 21,445 | -32% | 72 | 50 | -31% |
| White | 351,999 | 211,583 | -40% | 861 | 626 | -27% |
| Other | 6,880 | 4,455 | -35% | 21 | 24 | 14% |
| Unknown ethnicity | 53,411 | 48,525 | -9% | 91 | 96 | 5% |
| **Sexual orientation** |  |  |  |  |  |  |
| Heterosexual Males | 146,927 | 78,987 | -46% | 192 | 121 | -37% |
| MSM | 44,921 | 35,974 | -20% | 693 | 568 | -18% |
| Heterosexual Females | 283,049 | 185,791 | -34% | 220 | 154 | -30% |
| WSW | 2,866 | 2,767 | -3% | 3 | 3 | 0% |
| Unknown sexual orientation | 41,684 | 30,441 | -27% | 63 | 59 | -6% |
| **Area of residence** |  |  |  |  |  |  |
| Rural | 50,909 | 30,084 | -41% | 74 | 55 | -26% |
| Urban | 459,186 | 294,991 | -36% | 1,070 | 837 | -22% |
| Unknown area of residence | 9,352 | 8,885 | -5% | 27 | 13 | -52% |
| **Residential area-level deprivation (Deprivation Quintile)\*** |  |  |  |  |  |  |
| 1 (most deprived) | 114,267 | 70,837 | -38% | 357 | 275 | -23% |
| 2 | 128,123 | 84,643 | -34% | 327 | 245 | -25% |
| 3 | 102,827 | 66,940 | -35% | 219 | 168 | -23% |
| 4 | 87,324 | 54,707 | -37% | 149 | 111 | -26% |
| 5 (least deprived) | 77,554 | 47,948 | -38% | 92 | 93 | 1% |
| Unknown deprivation quintile | 9,352 | 8,885 | -5% | 27 | 13 | -52% |
| **Region of residence** |  |  |  |  |  |  |
| East Midlands | 38,033 | 19,792 | -48% | 72 | 43 | -40% |
| East of England | 48,082 | 28,046 | -42% | 59 | 59 | 0% |
| London | 142,416 | 103,316 | -27% | 382 | 315 | -18% |
| North–East | 20,684 | 11,430 | -45% | 91 | 78 | -14% |
| North–West | 55,936 | 30,863 | -45% | 205 | 127 | -38% |
| South–East | 82,258 | 56,890 | -31% | 137 | 99 | -28% |
| South–West | 45,034 | 28,029 | -38% | 61 | 63 | 3% |
| West Midlands | 42,700 | 23,298 | -45% | 76 | 60 | -21% |
| Yorkshire and Humber | 37,252 | 24,131 | -35% | 72 | 48 | -33% |
| Unknown region of residence | 7,052 | 8,165 | 16% | 16 | 13 | -19% |
| **HIV status** |  |  |  |  |  |  |
| HIV diagnosed | 949 | 618 | -35% | 58 | 43 | -26% |
| HIV negative or unknown | 518,498 | 333,342 | -36% | 1,113 | 862 | -23% |
| **Total** | **519,447** | **333,960** | **-36%** | **1,171** | **905** | **-23%** |

\*Deprivation quintile is an area-level measure of deprivation and socioeconomic status based on the 2019 Index of Multiple Deprivation (IMD) quintiles

**Table 3.** Number of chlamydia test and diagnoses among 15–24 year olds residing in England, by demographic characteristics: 2019–2020.

|  | Tests |  |  | Diagnoses |  |  |
| --- | --- | --- | --- | --- | --- | --- |
|  | 2019 | 2020 | Percentage difference | 2019 | 2020 | Percentage difference |
| **Age** |  |  |  |  |  |  |
| 15 to 19 | 418,019 | 258,419 | -38% | 48,138 | 31,140 | -35% |
| 20 to 24 | 914,914 | 670,879 | -27% | 80,288 | 57,165 | -29% |
| **Ethnicity** |  |  |  |  |  |  |
| Asian | 38,185 | 25,549 | -33% | 3,077 | 2,046 | -34% |
| Black African | 41,229 | 31,450 | -24% | 5,587 | 3,977 | -29% |
| Black Caribbean | 35,566 | 26,380 | -26% | 5,449 | 3,653 | -33% |
| Other Black ethnicity | 9,914 | 6,710 | -32% | 1,408 | 887 | -37% |
| Mixed ethnicity | 55,686 | 44,242 | -21% | 6,479 | 5,031 | -22% |
| White | 740,482 | 533,857 | -28% | 72,369 | 51,307 | -29% |
| Other | 11,440 | 7,974 | -30% | 1,145 | 794 | -31% |
| Unknown ethnicity | 400,431 | 253,136 | -37% | 32,912 | 20,610 | -37% |
| **Gender** |  |  |  |  |  |  |
| Female | 940,083 | 669,050 | -29% | 82,920 | 57,636 | -30% |
| Male | 380,647 | 252,121 | -34% | 44,173 | 29,476 | -33% |
| Unknown gender | 12,203 | 8,127 | -33% | 1,333 | 1,193 | -11% |
| **Online vs. Physical services** |  |  |  |  |  |  |
| Online services | 271,684 | 361,622 | 33% | 22,838 | 31,726 | 39% |
| Physical services | 1,041,553 | 554,299 | -47% | 104,343 | 55,607 | -47% |
| Unknown testing service | 19,696 | 13,377 | -32% | 1,245 | 972 | -22% |
| **Area of residence** |  |  |  |  |  |  |
| Rural | 147,884 | 106,001 | -28% | 12,899 | 9,017 | -30% |
| Urban | 1,105,688 | 774,434 | -30% | 107,641 | 74,482 | -31% |
| Unknown area of residence | 79,361 | 48,863 | -38% | 7,886 | 4,806 | -39% |
| **Residential area-level deprivation (Deprivation Quintile)\*** |  |  |  | 33,041 |  |  |
| 1 (most deprived) | 290,480 | 202,207 | -30% | 29,881 | 22,991 | -30% |
| 2 | 299,190 | 209,304 | -30% | 23,280 | 20,884 | -30% |
| 3 | 255,757 | 180,401 | -29% | 18,732 | 16,095 | -31% |
| 4 | 217,828 | 154,037 | -29% | 15,606 | 12,967 | -31% |
| 5 (least deprived) | 190,317 | 134,486 | -29% |  | 10,562 | -32% |
| Unknown deprivation quintile | 79,361 | 48,863 | -38% | 7,886 | 4,806 | -39% |
| **Region of residence** |  |  |  |  |  |  |
| East Midlands | 104,710 | 77,763 | -26% | 11,149 | 7,829 | -30% |
| East of England | 137,273 | 102,422 | -25% | 11,886 | 9,302 | -22% |
| London | 298,401 | 199,182 | -33% | 28,481 | 18,347 | -36% |
| North–East | 61,670 | 43,572 | -29% | 5,921 | 4,818 | -19% |
| North–West | 162,097 | 102,017 | -37% | 16,571 | 10,129 | -39% |
| South–East | 182,138 | 125,763 | -31% | 17,257 | 11,864 | -31% |
| South–West | 134,229 | 98,483 | -27% | 11,587 | 7,833 | 32% |
| West Midlands | 111,664 | 77,602 | -31% | 11,812 | 8,373 | -29% |
| Yorkshire and Humber | 140,751 | 102,494 | -27% | 13,762 | 9,810 | -29% |
| **Total** | **1,332,933** | **929,298** | **-30%** | **128,426** | **88,305** | **-31%** |

\*Deprivation quintile is an area-level measure of deprivation and socioeconomic status based on the 2019 Index of Multiple Deprivation (IMD) quintiles

Correlates of being tested for chlamydia via an online service Amongst all young people tested for chlamydia, those living in the least deprived areas were more likely to be tested online (unadjusted odds ratios - males: 1.24 [1.22-1.26]; females: 1.28 [1.27-1.30]) compared to young people living in the most deprived areas. This association remained after adjusting for confounders in the final model (males: 1.29 [1.27-1.32]; females 1.32 [1.30-1.34]) (Table 4). In the final model, there was a greater likelihood of being tested for chlamydia via an online service in 2020 [(males: 2.81 [2.77-2.84]; females: 2.45 [2.44-2.47]) vs. 2019] and a similarly increased likelihood amongst 20-24 year olds [(males: 1.47 [1.45-1.49]; females: 1.63 [1.61-1.64]) vs 15-19 year olds]. Online testing was also more likely among residents of urban areas [(males: 1.17 [1.15-1.20]; females: 1.16 [1.15-1.17]) vs rural] and was generally less likely among all regions of residence compared to London (*Appendix B*).

**Table 4.** Unadjusted and adjusted logistic regression analysis of the association between deprivation quintile^*^ and chlamydia testing via an online service among 15–24 year olds in England: 2019–2020, stratified by gender.

|  |  | Crude Odds Ratio<br>(95% CI) | Model 1, Adjusted<br>Odds Ratio**<br>(95% CI) | Model 2, Adjusted<br>Odds Ratio±<br>(95% CI) | Model 3, Adjusted<br>Odds Ratio¥<br>(95% CI) | Model 4, Adjusted<br>Odds Ratio‡<br>(95% CI) |
| --- | --- | --- | --- | --- | --- | --- |
| Male | **Residential area-level deprivation (Deprivation Quintile)\*** |  |  |  |  |  |
|  | 1 (most deprived) | 1 | – | – | – | – |
|  | 2 | 1.30 (1.28 – 1.33) | 1.32 (1.29 – 1.34) | 1.30 (1.28 – 1.32) | 1.31 (1.29 – 1.34) | 1.18 (1.16 – 1.20) |
|  | 3 | 1.40 (1.38 – 1.43) | 1.42 (1.40 – 1.45) | 1.41 (1.39 – 1.43) | 1.46 (1.43 – 1.48) | 1.35 (1.32 – 1.37) |
|  | 4 | 1.36 (1.34 – 1.39) | 1.39 (1.36 – 1.41) | 1.39 (1.36 – 1.41) | 1.45 (1.42 – 1.48) | 1.37 (1.35 – 1.40) |
|  | 5 (least deprived) | 1.24 (1.22 – 1.26) | 1.25 (1.23 – 1.28) | 1.26 (1.23 – 1.28) | 1.31 (1.29 – 1.34) | 1.29 (1.27 – 1.32) |
| Female | **Residential area-level deprivation (Deprivation Quintile)\*** |  |  |  |  |  |
|  | 1 (most deprived) | 1 | – | – | – | – |
|  | 2 | 1.34 (1.32 – 1.35) | 1.35 (1.34 – 1.37) | 1.34 (1.32 – 1.35) | 1.35 (1.34 – 1.37) | 1.20 (1.19 – 1.21) |
|  | 3 | 1.40 (1.39 – 1.42) | 1.42 (1.40 – 1.43) | 1.40 (1.39 – 1.42) | 1.46 (1.44 – 1.47) | 1.33 (1.32 – 1.35) |
|  | 4 | 1.39 (1.38 – 1.41) | 1.40 (1.39 – 1.42) | 1.40 (1.38 – 1.41) | 1.46 (1.45 – 1.48) | 1.38 (1.36 – 1.39) |
|  | 5 (least deprived) | 1.28 (1.27 – 1.30) | 1.29 (1.27 – 1.30) | 1.29 (1.28 – 1.31) | 1.35 (1.33 – 1.37) | 1.32 (1.30 – 1.34) |

\*Deprivation quintile is an area-level measure of deprivation and socioeconomic status based on the 2019 Index of Multiple Deprivation (IMD) quintiles \*\* Model 1 adjusted for year of test ± Model 2 adjusted for year of test and age group ¥ Model 3 adjusted for year of test, age group and area of residence ‡ Model 4 adjusted for year of test, age group, area of residence and region

## Discussion

There was a decrease in STI testing and diagnoses among young people during the first year of the COVID-19 pandemic, with up to 50% larger decreases in teenagers. In keeping with the reconfiguration of SHSs in 2020 to offer more remote consultations, we found a 33% increase in chlamydia testing of young people via online services, but there was evidence of inequalities in access to testing via this modality.

Among young people tested for chlamydia, those living in the least deprived areas were more likely to be tested for chlamydia online, compared to those living in the most deprived areas. Further inequalities in chlamydia online testing were found, with young people living in rural areas or regions outside London and those aged 15-19 being less likely to be tested for chlamydia using an online service. This suggests that there may be socioeconomic or structural barriers to online testing, which may include lack of online access. 15-19 year olds may find it more difficult to be tested for chlamydia using an online service if they are still living with their parents and are unable to discreetly receive the self-sampling kit. The greater likelihood of being tested for chlamydia online for young people living in London reflects the fact that there is a pan-London online sexual health service for all London residents^[24]^.

The reductions in STI testing between 2019 and 2020 are partly due to the extensive public health measures to help reduce the transmission of COVID-19^[25]^. Moreover, individuals may have delayed their visits to SHSs due to fear of COVID infection^[26]^ and with lockdown restrictions it would have been difficult to meet and interact with new people, reducing the possibility of new sexual encounters^[26]^. All these factors may have contributed to the decline in STI testing in 2020.

Our findings are consistent with international literature highlighting the negative impact the COVID-19 pandemic had on STI testing. A report from the EuroTEST COVID-19 impact assessment consortium found that, among 34 countries in the World Health Organization European Region and in different test settings, 95% of them tested less than half the expected number of people between March and May 2020, this decline continued until August 2020^[27]^. Research in the USA found a reduction in STI testing and case detection resulting in more than, 27,000 missed cases of chlamydia and 5,500 cases of gonorrhoea between March and June 2020^[25]^. The implications of these missed cases are likely to be increased community transmission due to the asymptomatic nature of these STIs and associated long-term sexual and reproductive health complications^[25]^. Studies have found that testing via online services is preferred over physical services, particularly amongst young people^[28]^, but this may not be the case for teenagers who are living at home. The advantages of online services include privacy and the ability to self-sample^[29]^. Previous research has found that online testing is more likely to be used by women and those between the ages of 20-30 compared to younger age groups. Consistent with our findings, research conducted amongst online services and SHSs in London found those living in less deprived areas are more likely to use online services when testing for an STI even when adjusting for confounders^[30]^.

Our analysis benefitted from a very large sample from national surveillance datasets which included patient-level data with key demographic factors so we could robustly assess differences in testing patterns within different subgroups. However, our analysis is not without limitations. Urban and rural area classifications were based on the 2011 census (the most up to date dataset at the time of writing) and these may not be accurate for all areas of England in 2020. We were unable to adjust for ethnicity in the regression analysis predicting being tested online for chlamydia due to a high proportion of missing values for ethnicity in the CTAD surveillance system. The regression analysis was restricted to chlamydia because we were only able to reliably identify all sources of online testing for chlamydia by using a combination of CTAD and GUMCAD data at the time of writing. While GUMCAD is a rich source of data on STI testing, it underestimates online testing for gonorrhoea and syphilis as it could only identify online testing by standalone online providers, and not online testing provided as an alternate service by physical SHSs, in 2019 and 2020.

Similarly, we did not perform a regression analysis with count data to determine correlates of being tested for chlamydia online vs not being tested – this is because of a lack of underlying population data for all key variables (e.g. age-group, gender and residential area-level deprivation). However, as we have comprehensive data on all young people tested for chlamydia, we were able to assess the correlates of being tested online. Whilst we included deprivation quintile in our analyses, this in an area-level, rather than individual-level, measure of deprivation and is subject to the ecologic fallacy. Additionally, the larger proportional drop in STI tests among teenagers may be explained by residual confounding as our analyses could not take risk behaviours such as multiple condomless sex partners into account, and it is unclear how this varied between 15-19 and 20-24 year olds between 2019 and 2020.

Reduced testing, missed infections and late diagnoses may have potential consequences such as the increase in PID and infertility^[31]^. This will impact the quality of life of young people with STIs and increase costs to the healthcare system with the need for treatments for STI-related complications or sequelae. Additionally, the difference in the means of testing between those in the least and most deprived areas suggests barriers to access to online services, which should not occur, as STI testing is free at the point of delivery in England. Given the increasing shift to online service provision, there remains a need to assess how equitably they are provided and to reduce the risk of differential access widening existing inequalities in sexual health.

## Data Availability

All data produced in the present work are contained in the manuscript. Further data can be found on the UKHSA website.

https://www.gov.uk/government/statistics/sexually-transmitted-infections-stis-annual-data-tables

https://www.gov.uk/government/statistics/national-chlamydia-screening-programme-ncsp-data-tables

## Acknowledgments

The authors thank all laboratories and sexual health services that report CTAD and GUMCAD surveillance data to UKHSA. We also thank colleagues in the CTAD, GUMCAD and NCSP teams at UKHSA. We acknowledge members of the National Institute for Health Research Health Protection Research Unit (NIHR HPRU) in Blood Borne and Sexually Transmitted Infections (BBSTI) Steering Committee: Professor Caroline Sabin (HPRU Director), Dr John Saunders (UKHSA Lead), Professor Catherine Mercer, Dr Hamish Mohammed (previously Professor Gwenda Hughes), Professor Greta Rait, Dr Ruth Simmons, Professor William Rosenberg, Dr Tamyo Mbisa, Professor Rosalind Raine, Dr Sema Mandal, Dr Rosamund Yu, Dr Samreen Ijaz, Dr Fabiana Lorencatto, Dr Rachel Hunter, Dr Kirsty Foster and Dr Mamooma Tahir.

## Appendix A

**Table A1).** Demographic characteristics of 15–24 year old males tested for chlamydia in England: 2019–2020

|  | Number and proportion tested<br>in online services | Number and proportion tested<br>in physical services |
| --- | --- | --- |
| **Age group (years)** |  |  |
| 15–19 | 37,627<br>(23.1%) | 124,890<br>(76.8%) |
| 20–24 | 152,859<br>(32.5%) | 317,392<br>(67.5%) |
| **Area of residence** |  |  |
| Rural | 20,521<br>(28.2%) | 52,145<br>(71.8%) |
| Urban | 165,813<br>(89.0%) | 356,975<br>(68.3%) |
| Unknown area of residence | 4,152<br>(11.1%) | 33,162<br>(88.9%) |
| **Region of residence** |  |  |
| London | 58,322<br>(37.0%) | 99,114<br>(63.0%) |
| East Midlands | 18,518<br>(38.5%) | 29,578<br>(61.5%) |
| East of England | 17,513<br>(25.2%) | 52,085<br>(74.8%) |
| North–East | 5,240<br>(18.9%) | 22,510<br>(81.1%) |
| North–West | 10,012<br>(14.6%) | 58,466<br>(85.4%) |
| South–East | 21,454<br>(25.7%) | 61,869<br>(74.2%) |
| South–West | 22,052<br>(34.6%) | 41,587<br>(65.3%) |
| West Midlands | 14,520<br>(27.8%) | 37,618<br>(72.1%) |
| Yorkshire and Humber | 22,855<br>(36.7%) | 39,455<br>(63.3%) |
| **Year of test** |  |  |
| 2019 | 81,880<br>(21.5%) | 298,767<br>(78.5%) |
| 2020 | 108,606<br>(43.1%) | 143,515<br>(56.9%) |
| **Residential area-level deprivation**<br>**(Deprivation Quintile)\*** |  |  |
| 1 (most deprived) | 34,679<br>(26.6%) | 95,436<br>(73.3%) |
| 2 | 45,911<br>(32.1%) | 96,871<br>(67.8%) |
| 3 | 41,352<br>(33.8%) | 81,030<br>(66.2%) |
| 4 | 35,331<br>(33.0%) | 71,259<br>(66.8%) |
| 5 (least deprived) | 29,061<br>(31.3%) | 64,524<br>(68.7%) |
| Unknown deprivation quintile | 4,152<br>(11.1%) | 33,162<br>(88.9%) |
| **Total** | **190,486** | **442,282** |

**Table A2).** Demographic characteristics of 15–24 year old females tested for chlamydia in England: 2019–2020

|  | Number and proportion tested<br>in online services | Number and proportion tested<br>in physical services |
| --- | --- | --- |
| **Age group (years)** |  |  |
| 15–19 | 102,232<br>(20.2%) | 404,106<br>(79.8%) |
| 20–24 | 337,692<br>(30.6%) | 765,103<br>(69.4%) |
| **Area of residence** |  |  |
| Rural | 44,744<br>(24.9%) | 135,043<br>(75.1%) |
| Urban | 383,826<br>(28.6%) | 957,935<br>(71.4%) |
| Unknown area of residence | 11,354<br>(13.0%) | 76,231<br>(87.0%) |
| **Region of residence** |  |  |
| London | 123,797<br>(36.8%) | 212,252<br>(63.2%) |
| East Midlands | 43,825<br>(32.8%) | 89,923<br>(67.2%) |
| East of England | 38,838<br>(22.9%) | 130,417<br>(77.0%) |
| North–East | 11,834<br>(15.7%) | 63,595<br>(84.3%) |
| North–West | 29,550<br>(15.4%) | 162,180<br>(84.6%) |
| South–East | 52,636<br>(24.0%) | 166,789<br>(76.0%) |
| South–West | 50,939<br>(30.2%) | 117,440<br>(69.7%) |
| West Midlands | 33,625<br>(24.9%) | 101,346<br>(75.1%) |
| Yorkshire and Humber | 54,880<br>(30.5%) | 125,267<br>(30.5%) |
| **Year of test** |  |  |
| 2019 | 188,169<br>(20.0%) | 751,914<br>(80.0%) |
| 2020 | 251,755<br>(37.6%) | 417,295<br>(62.4%) |
| **Residential area-level deprivation**<br>**(Deprivation Quintile)\*** |  |  |
| 1 (most deprived) | 84,665<br>(23.6%) | 273,626<br>(76.4) |
| 2 | 105,764<br>(29.3%) | 255,330<br>(70.7) |
| 3 | 94,142<br>(30.3%) | 216,725<br>(69.7%) |
| 4 | 79,044<br>(30.1%) | 183,472<br>(69.9%) |
| 5 (least deprived) | 64,955<br>(28.4%) | 163,825<br>(71.6%) |
| Unknown deprivation quintile | 11,354<br>(13.0%) | 76,231<br>(87.0%) |
| **Total** | **439,924** | **1,169,209** |

\*Deprivation quintile is an area-level measure of deprivation and socioeconomic status based on the 2019 Index of Multiple Deprivation (IMD) quintiles

## Appendix B

**Table B1).** Adjusted logistic regression analysis of the association between deprivation quintile^*^ and chlamydia testing via an online service among 15–24 year old males in England: 2019–2020

|  | Adjusted Odds Ratio (95% CI) |
| --- | --- |
| **Age group (years)** |  |
| 15–19 | 1 |
| 20–24 | 1.47 (1.45–1.49) |
| **Area** |  |
| Rural | 1 |
| Urban | 1.17 (1.15–1.20) |
| **Region of residence** |  |
| London | 1 |
| East Midlands | 1.03 (1.01–1.05) |
| East of England | 0.49 (0.48–0.50) |
| North–East | 0.37 (0.35–0.38) |
| North–West | 0.26 (0.25–0.26) |
| South–East | 0.56 (0.55–0.58) |
| South–West | 0.84 (0.82–0.85) |
| West Midlands | 0.62 (0.60–0.63) |
| Yorkshire and Humber | 0.93 (0.91–0.95) |
| **Year of test** |  |
| 2019 | 1 |
| 2020 | 2.81 (2.77–2.84) |
| **Residential area-level deprivation (Deprivation Quintile)\*** |  |
| 1 (most deprived) | 1 |
| 2 | 1.18 (1.16 –1.20) |
| 3 | 1.35 (1.32–1.37) |
| 4 | 1.37 (1.35–1.40) |
| 5 (least deprived) | 1.29 (1.27–1.32) |

\*Deprivation quintile is an area-level measure of deprivation and socioeconomic status based on the 2019 Index of Multiple Deprivation (IMD) quintiles
\*\* All p-values <0.05

**Table B2).** Adjusted logistic regression analysis of the association between deprivation quintile^*^ and chlamydia testing via an online service among 15–24 year old females in England: 2019–2020

|  | Adjusted Odds Ratio (95% CI) |
| --- | --- |
| **Age group (years)** |  |
| 15–19 | 1 |
| 20–24 | 1.63 (1.61–1.64) |
| **Area** |  |
| Rural | 1 |
| Urban | 1.16 (1.15–1.17) |
| **Region of residence** |  |
| London | 1 |
| East Midlands | 0.80 (0.79–0.81) |
| East of England | 0.44 (0.43–0.44) |
| North–East | 0.31 (0.30–0.31) |
| North–West | 0.28 (0.27–0.28) |
| South–East | 0.51 (0.50–0.51) |
| South–West | 0.68 (0.67–0.69) |
| West Midlands | 0.53 (0.52–0.54) |
| Yorkshire and Humber | 0.71 (0.70–0.72) |
| **Year of test** |  |
| 2019 | 1 |
| 2020 | 2.45 (2.44–2.47) |
| **Residential area-level deprivation (Deprivation Quintile)\*** |  |
| 1 (most deprived) | 1 |
| 2 | 1.20 (1.19 –1.21) |
| 3 | 1.33 (1.32–1.35) |
| 4 | 1.38 (1.36–1.39) |
| 5 (least deprived) | 1.32 (1.30–1.34) |

\*Deprivation quintile is an area-level measure of deprivation and socioeconomic status based on the 2019 Index of Multiple Deprivation (IMD) quintiles
\*\* All p-values <0.05

## Appendix C

**Table C1).**
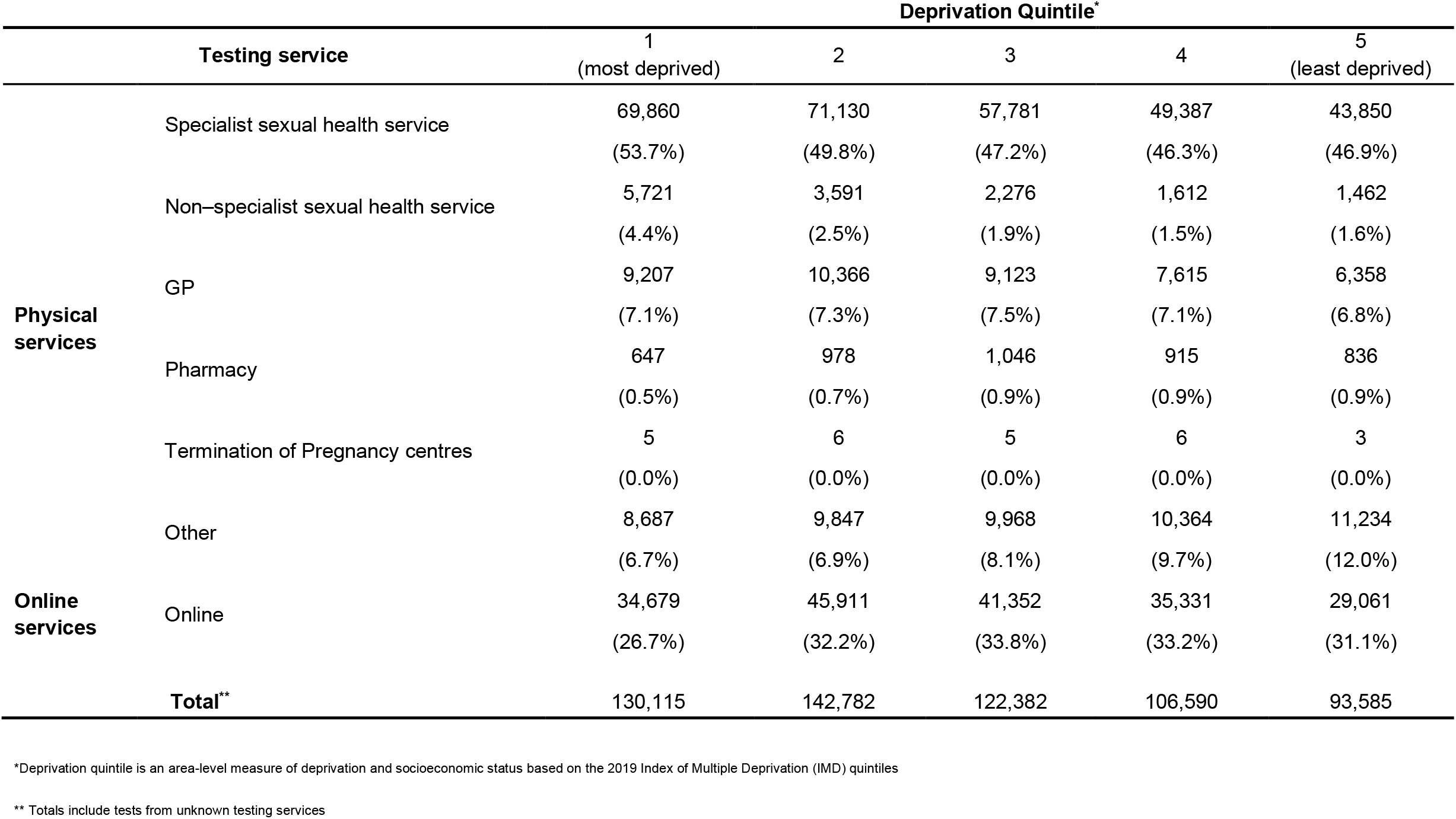
Number and proportion of chlamydia tests by testing service and deprivation quintile^*^ amongst 15–24 year old males in England: 2019–2020

**Table C2).** Number and proportion of chlamydia tests by testing service and deprivation quintile^*^ amongst 15–24 year old females in England: 2019–2020

|  |  | Deprivation Quintile* |  |  |  |  |
| --- | --- | --- | --- | --- | --- | --- |
| Testing service |  | 1<br>(most deprived) | 2 | 3 | 4 | 5<br>(least deprived) |
| **Physical services** | Specialist sexual health service | 138,082<br>(38.5%) | 133,456<br>(37.0%) | 107,714<br>(34.6%) | 89,520<br>(34.1%) | 78,302<br>(34.2%) |
|  | Non-specialist sexual health service | 16,847<br>(4.7%) | 11,184<br>(3.1%) | 6,816<br>(2.2%) | 4,893<br>(1.9%) | 3,792<br>(1.7%) |
|  | GP | 67,115<br>(18.7%) | 64,873<br>(18.0%) | 60,091<br>(19.3%) | 53,046<br>(20.2%) | 49,186<br>(21.5%) |
|  | Pharmacy | 1,834<br>(0.5%) | 2,653<br>(0.7%) | 3,063<br>(1.0%) | 2,598<br>(1.0%) | 2,561<br>(1.1%) |
|  | Termination of Pregnancy centres | 8,325<br>(2.3%) | 7,248<br>(2.0%) | 5,142<br>(1.7%) | 3,708<br>(1.4%) | 2,971<br>(1.3%) |
|  | Other | 35,464<br>(9.9%) | 31,428<br>(8.7%) | 29,881<br>(9.6%) | 24,528<br>(9.3%) | 23,889<br>(10.4%) |
|  | **Online services** |  |  |  |  |  |
|  | Online | 84,665<br>(23.6%) | 105,764<br>(29.3%) | 94,142<br>(30.3%) | 79,044<br>(30.1%) | 64,955<br>(28.4%) |
| **Total\*\*** |  | 358,291 | 361,094 | 310,867 | 262,516 | 228,780 |

\*Deprivation quintile is an area-level measure of deprivation and socioeconomic status based on the 2019 Index of Multiple Deprivation (IMD) quintiles
\*\* Totals include tests from unknown testing services

## References

[1] Mapp F, Hickson F, Mercer CH, Wellings K. How social representations of sexually transmitted infections influence experiences of genito-urinary symptoms and care-seeking in Britain: mixed methods study protocol. BMC public health, 2016. 16(1): p. 1–9. DOI: 10.1186/s12889-016-3261-0.

[2] Clifton S, Mercer CH, Sonnenberg P, et al STI risk perception in the British Population and how it relates to sexual Behaviour and STI Healthcare use: findings from a cross-sectional Survey (Natsal-3). EClinicalMedicine, 2018. 2: p. 29–36. DOI: 10.1016/j.eclinm.2018.08.001.

[3] Mitchell H, Allen H, Sonubi T, et al. Sexually transmitted infections and screening for chlamydia in England, 2019. Public Health England (PHE), London. 2020.

[4] Mercer CH, Tanton C, Prah P, et al. Changes in sexual attitudes and lifestyles in Britain through the life course and over time: findings from the National Surveys of Sexual Attitudes and Lifestyles (Natsal). The Lancet, 2013. 382(9907): p. 1781–1794. DOI: 10.1016/S0140-6736(13)62035-8.

[5] Sonnenberg P, Menezes D, Freeman L, et al. Intimate physical contact between people from different households during the COVID-19 pandemic: a mixed-methods study from a large, quasi-representative survey (Natsal-COVID). BMJ open, 2022. 12(2): p. e055284. DOI: 10.1136/bmjopen-2021-055284.

[6] Tanton C, Geary RS, Clifton S, et al. Sexual health clinic attendance and non-attendance in Britain: findings from the third National Survey of Sexual Attitudes and Lifestyles (Natsal-3). Sexually Transmitted Infections, 2018. 94(4): p. 268–276. DOI: 10.1136/sextrans-2017-053193.

[7] Sonnenberg P, Clifton S, Beddows S, et al. Prevalence, risk factors, and uptake of interventions for sexually transmitted infections in Britain: findings from the National Surveys of Sexual Attitudes and Lifestyles (Natsal). The Lancet. 2013. 382. DOI: 10.1016/S0140-6736(13)61947-9.

[8] Tiller, CM. Chlamydia during pregnancy: implications and impact on perinatal and neonatal outcomes. Journal of Obstetric, Gynecologic, & Neonatal Nursing, 2002. 31(1): p. 93–98. DOI: 10.1111/j.1552-6909.2002.tb00027.x.

[9] Centers for Disease Control and Prevention (CDC). and Prevention, Sexually transmitted disease surveillance 2012. Centers for Disease Control and Prevention, Atlanta, GA. 2014.

[10] Detels R, Green AM, Klausner JD, et al. The incidence and correlates of symptomatic and asymptomatic Chlamydia trachomatis and Neisseria gonorrhoeae infections in selected populations in five countries. Sexually transmitted diseases, 2011. 38(6): p. 503.

[11] JoãO AL, Lencastre A, CalvãO J, et al. COVID-19, fear and sexual behaviour: a survey in a tertiary STI clinic in Lisbon. Sexually Transmitted Infections, 2021. 97(7): p. 549–549. DOI: 10.1136/sextrans-2020-054834.

[12] Bardsley M, Wayal S, Blomquist P, et al. Improving our understanding of the disproportionate incidence of STIs in heterosexual-identifying people of black Caribbean heritage: findings from a longitudinal study of sexual health clinic attendees in England. Sexually Transmitted Infections, 2022. 98(1): p. 23–31. DOI: 10.1136/sextrans-2020-054784.

[13] Savage EJ, Mohammed H, Leong G, et al. Improving surveillance of sexually transmitted infections using mandatory electronic clinical reporting: the genitourinary medicine clinic activity dataset, England, 2009 to 2013. Euro Surveillance, 2014. 19(48): p. 20981. DOI: 10.2807/1560-7917.es2014.19.48.20981.

[14] Public Health England (PHE). Chlamydia Testing Activity Dataset: Dataset specification and technical guidance. Public Health England, London. 2016.

[15] Chandra NL, Soldan K, Dangerfield C, et al. Filling in the gaps: estimating numbers of chlamydia tests and diagnoses by age group and sex before and during the implementation of the English National Screening Programme, 2000 to 2012. Euro Surveillance, 2017. 22(5): p. 23–33. DOI: 10.2807/1560-7917.ES.2017.22.5.30453.

[16] UK Health Security Agency (UKHSA). GUMCAD STI Surveillance System. Data specification and technical guidance. UK Health Security Agency (UKHSA), London 2021.

[17] Office for National Statistics (ONS). Super Output Area (SOA). Available at: https://webarchive.nationalarchives.gov.uk/ukgwa/20160106001702/http://www.ons.gov.uk/ons/guide-method/geography/beginner-s-guide/census/super-output-areas--soas-/index.html [accessed 13.07.22].

[18] Ministry of Housing, Communities and Local Government (MHCLG). The English Indices of Deprivation 2019 (IoD2019). 2019.

[19] Office for National Statistics (ONS). 2011 Census: Characteristics of Built-Up Areas. 2013. Available at: https://www.ons.gov.uk/peoplepopulationandcommunity/housing/articles/characteristicsofbuiltupareas/2013-06-28 [accessed 12.05.22].

[20] Office for National Statistics (ONS). 2011 Built-up Areas - Methodology and Guidance. Office for National Statistics: London. 2013.

[21] Office for National Statistics (ONS). 2011 rural/urban classification. 2016. Available at: https://www.ons.gov.uk/methodology/geography/geographicalproducts/ruralurbanclassifications/2011ruralurbanclassification [accessed: 12.05.22].

[22] Office for National Statistics (ONS). Ethnic group, national identity and religion. Available at: https://www.ons.gov.uk/methodology/classificationsandstandards/measuringequality/ethnicgroupnationalidentityandreligion [accessed 07.07.22].

[23] Statacorp, LP., Stata data analysis and statistical Software. Special Edition Release, 2007. 10: p. 733.

[24] Sexual Health London (SHL). Home STI testing, regular and emergency contraception. Free NHS sexual health services online. Available at: https://www.shl.uk/ [accessed 20.07.22].

[25] Pinto CN, Niles JK, Kaufman HW, et al. Impact of the COVID-19 Pandemic on Chlamydia and Gonorrhea Screening in the US. American journal of preventive medicine, 2021. 61(3): p. 386–393. DOI: 10.1016/j.amepre.2021.03.009.

[26] Latini A, Magri F, Dona MG, et al. Is COVID-19 affecting the epidemiology of STIs? The experience of syphilis in Rome. Sexually Transmitted Infections, 2021. 97(1): p. 78. DOI: 10.1136/sextrans-2020-054543.

[27] SimõEs D, Stengaard AR, Combs L, Rabenet D. Impact of the COVID-19 pandemic on testing services for HIV, viral hepatitis and sexually transmitted infections in the WHO European Region, March to August 2020. Eurosurveillance, 2020. 25(47): p. 2001943. DOI: 10.2807/1560-7917.ES.2020.25.47.2001943.

[28] Eaton S, Biggerstaff D, Petrou S, et al. Young people’s preferences for the use of emerging technologies for asymptomatic regular chlamydia testing and management: a discrete choice experiment in England. BMJ Open, 2019. 9(1). DOI: 10.1136/bmjopen-2018-023663.

[29] Spence T, Kander I, Walsh J, et al. Perceptions and Experiences of Internet-Based Testing for Sexually Transmitted Infections: Systematic Review and Synthesis of Qualitative Research. Journal of Medical Internet Research, 2020. 22(8). DOI: 10.2196/17667.

[30] Barnard S, Free C, Bakolis I, et al. Comparing the characteristics of users of an online service for STI self-sampling with clinic service users: a cross-sectional analysis. Sexually Transmitted Infections, 2018. 94(5): p. 377–383. DOI: 10.1136/sextrans-2017-053302.

[31] O’Connell CM, Ferone ME., Chlamydia trachomatis Genital Infections. Microbial Cell, 2016. 3(9): p. 390–403. DOI: 10.15698/mic2016.09.525.

